# Positive effects of functional electrical stimulation-assisted cycling on perception of effort, cerebral blood flow and cognition in post-stroke patients

**DOI:** 10.1101/2025.06.24.25330073

**Authors:** Maël Descollonges, Julie Di Marco, Ehsan Jafari, Pierre-Henri Pouillart, Julien V. Brugniaux, Benjamin Pageaux, Gaëlle Deley

**Affiliations:** INSERM UMR 1093 – Laboratoire CAPS, Université de Bourgogne, UFR des Sciences du Sport, Dijon, France; Kurage, Lyon, France; SMR Val Rosay, Saint-Didier au Mont d’Or, France; Université de Lyon, ENS de Lyon, CNRS, Laboratoire de Physique, F-69342 Lyon, France; HP2 laboratory, Univ. Grenoble Alpes, INSERM, CHU Grenoble Alpes, Grenoble, France; Centre de recherche de l’Institut universitaire de gériatrie de Montréal (CRIUGM), Canada; École de kinésiologie et des sciences de l’activité physique (EKSAP), Faculté de médecine, Université de Montréal, Montréal, Canada; Centre Interdisciplinaire de Recherche sur le Cerveau et l’Apprentissage (CIRCA), Montréal, QC, Canada

**Keywords:** Cerebral blood flow, cognition, functional electrical stimulation, rating of perceived exertion, stroke

## Abstract

**Background:** FES-assisted cycling may reduce perceived effort by lowering the required motor command compared to voluntary cycling. While benefits on effort perception have been shown during walking in stroke and multiple sclerosis patients, its effectiveness during cycling in stroke rehabilitation remains unproven. Thus, this work aimed to test the effect of functional electrical stimulation-assisted cycling on stroke patients’ perception of effort (primary aim) in a randomized controlled study design. In an exploratory way, this work also aimed to examine the effect of FES-assisted cycling on cerebral blood flow and cognitive performance (exploratory aims) in a subsample.

**Methods:** Fifteen post-stroke patients completed functional electrical stimulation-assisted and traditional cycling sessions separated by 72h. Perceived effort, cardio, and cerebrovascular parameters were monitored during exercise. Cognitive performance was assessed before and after each session. Qualitative data were reported after both sessions.

**Results:** Patients reported a lower perceived effort during functional electrical stimulation-assisted cycling than traditional cycling. Both sessions increased heart rate, end-tidal CO_2_, cardiac output, and cerebral artery blood flow velocity, with higher blood lactate levels after functional electrical stimulation-assisted cycling. Functional electrical stimulation-assisted cycling positively impacted cognitive performance.

**Conclusion:** Traditional and functional electrical stimulation-assisted cycling induced similar increased cardio and cerebrovascular responses. However, patients perceived functional electrical stimulation-assisted cycling as less effortful than traditional cycling. As effort is a barrier to regular exercise engagement and adherence, these results are promising for implementing functional electrical stimulation-assisted cycling in stroke patients’ rehabilitation.

## Background

Stroke is a major cause of long-term disability affecting walking, postural control, muscle strength, cerebrovascular and cardiovascular systems, and cognition. To counteract cardiorespiratory system deconditioning and experience the positive effect of physical exercise on cognition (1), stroke patients must engage in regular aerobic exercise during their rehabilitation. Due to hemiparesis and fatigue leading to increased effort perception (2), and the proposed key role of effort and its perception in the engagement in physical and cognitive activities (3), post-stroke individuals experience difficulty engaging in and performing physical exercise. Therefore, developing and implementing methods of rehabilitation that minimize the perception of effort experienced during physical exercise is crucial.

To this aim, functional electrical stimulation (FES)-assisted cycling is a promising method. Clinical evidence shows that FES-assisted cycling activates paralyzed muscles (4) and improves aerobic capacity and cardiopulmonary function (5). By evoking muscle contraction superimposed on top of the impaired voluntary contractions, FES is known to assist in movement execution (4). As the evoked contraction induced by FES pre-activates the muscles, the individual engaged in FES-assisted cycling must voluntarily produce a lower muscle force to reach the targeted exercise intensity. As effort perception is proposed to be generated by signals related to the magnitude of the motor command (6,7) and the lower muscle force to be voluntarily produced during FES should be associated with a lower motor command compared to traditional cycling, using FES-assisted cycling is a promising tool to decrease the level of perceived effort experienced during rehabilitation sessions. To date, it has been reported that FES application was associated with a reduced perception of effort in stroke or multiple sclerosis patients during walking tasks (8,9). However, the effectiveness in reducing perceived effort during cycling in stroke patients remains to be validated. As cycling is a key aerobic exercise in rehabilitation, there is an urgent need for such a study.

In this context, this study aimed to compare the perceived effort during an FES-assisted cycling session, compared to a traditional cycling session, in stroke patients. We hypothesized that stroke patients would perceive a lower effort during FES-assisted cycling compared to traditional cycling, performed at the same workload. In addition, considering recent research suggesting a positive effect of electrical muscle stimulation on cerebral blood flow velocity (CBFv, (10,11)) and potentially cognitive performance, we also explored, in a subsample of patients, changes in CBFv to the FES- and traditional cycling exercises, and cognitive performance pre-post exercises.

## Methods

Fifteen post-stroke patients volunteered in this experiment (8♀; 7♂; age: 53 ± 13 years, height: 167 ± 0 cm, mass: 69 ± 10 kg, body mass index: 25 ± 3, 148 ± 108 days after stroke). Patients’ characteristics stratified by sex are available in Table 1.

**Table 1:** Patient’s characteristics. I: Ischemic; H: Hemorrhagic.

| Females |  |  |  |  |  |  |  |  |
| --- | --- | --- | --- | --- | --- | --- | --- | --- |
| Patients | Age (years) | Size (cm) | Weight (kg) | BMI | Type of stroke | Days after stroke | Hemiparetic side | Assistive device |
| 1 | 35-40 | 150 | 67 | 29.8 | H | 189 | Left | AFO |
| 2 | 40-45 | 169 | 69 | 24.2 | I | 40 | Right | - |
| 3 | 75-80 | 170 | 64 | 22.1 | I | 90 | Right | AFO |
| 4 | 50-55 | 155 | 63 | 26.1 | I | 90 | Right | Cane |
| 5 | 35-40 | 160 | 79 | 30.9 | I | 80 | Right | Cane |
| 6 | 35-40 | 170 | 74 | 25.6 | H | 237 | Right | AFO |
| 7 | 55-60 | 158 | 48 | 19.2 | I | 385 | Left | AFO |
| 8 | 65-70 | 163 | 68 | 25.5 | I | 213 | Left | Wheelchair |
| Mean | 51 | 160 | 66 | 25 | - | 166 | - | - |
| Males |  |  |  |  |  |  |  |  |
| 9 | 55-60 | 175 | 67 | 21.9 | I | 330 | Left | AFO |
| 10 | 65-70 | 165 | 66 | 24.2 | I | 38 | Left | AFO |
| 11 | 60-65 | 175 | 78 | 25.5 | I | 90 | Left | - |
| 12 | 35-40 | 162 | 74 | 28.1 | H | 97 | Left | Cane |
| 13 | 55-60 | 167 | 66 | 23.7 | I | 104 | Right | - |
| 14 | 45-50 | 186 | 91 | 26,4 | H | 207 | Left | Wheelchair + cane |
| 15 | 50-55 | 180 | 60 | 20.1 | I | 33 | Right | - |
| Mean | 55 | 173 | 72 | 24 | - | 128 | - | - |

Each patient provided written informed consent describing the procedure and potential risks of the study. The sample size was based on patients’ availability at the time of the experiment and an expected large effect size of FES-assisted cycling on the perception of effort. Inclusion criteria were diagnosis of an ischemic (n=11) or hemorrhagic (n=4) stroke resulting in hemiparesis; adequate cognitive abilities to engage in rehabilitation activities; able to sit up to 30 minutes; joint mobility ranges that would not preclude pedaling. The study was supported by UGECAM Rhône-Alpes and received ethical approval from the Comité de Protection des Personnes Sud-Est II (French Institutional Review Board). The study was approved under the clinical trial registration number NCT06230796.

Patients visited the hospital on six occasions. In visit 1, participants performed an incremental test to determine their peak power output (PPO). The incremental test started at 25 watts, with the power output increasing by 10 watts every minute until exhaustion. Exhaustion was defined as voluntary disengagement from the task or an inability to maintain a cadence above 50 revolutions/minute (rpm) for more than 3 seconds, despite verbal encouragement. In visits 2-3- 4, patients were extensively familiarized with FES, the cognitive tasks, and the psychophysical scale used to report their perceived effort. Each familiarization visit was separated by 48 hours. In visits 5-6, participants performed two testing sessions in a randomized order: FES-assisted cycling and traditional cycling. Testing sessions were separated by 72 hours to control for the presence of muscle fatigue and soreness in the second testing session (visit 6).

During FES-assisted cycling, electrical stimulation was applied to the quadriceps, hamstrings, and gluteus muscles of the hemiparesis side during cycling using a stimulator (Motimove, 3F- Fabricando, Belgrade, Serbia) and an electrical stimulation-assisted cycle ergometer (Kurage, HephaBike, Lyon, France) (Figure 1). The paretic leg was secured in alignment using an orthotic pedal, ensuring stabilization of the foot and leg during the exercise (Figure 1). A computerized program linked to an encoder fixed in the pedal axis was employed to stimulate the muscles in the movement’s proper phase. The appropriate phase corresponds to the knee extension and knee flexion (pedaling movement). Quadriceps and gluteus muscles were stimulated during extension, and hamstrings during flexion. The stimulation frequency was fixed at 40 Hz, with a pulse width of 400 μs, and the stimulation intensity (mA) was constantly adjusted to the individual maximum tolerable level throughout the session. The maximum tolerable intensity of stimulation was determined by gradually increasing the current intensity until an effective contraction was achieved and further increased to elicit the maximal pain intensity tolerated by the participant. Each session began with a 5-minute warm-up consisting of a progressive increase in stimulation intensity and cycling resistance to reach 50% PPO. Afterwards, the resistance was fixed at 50% PPO for 20 minutes, and patients had to maintain a cadence of 50 ± 3 rpm.

**Figure 1:**
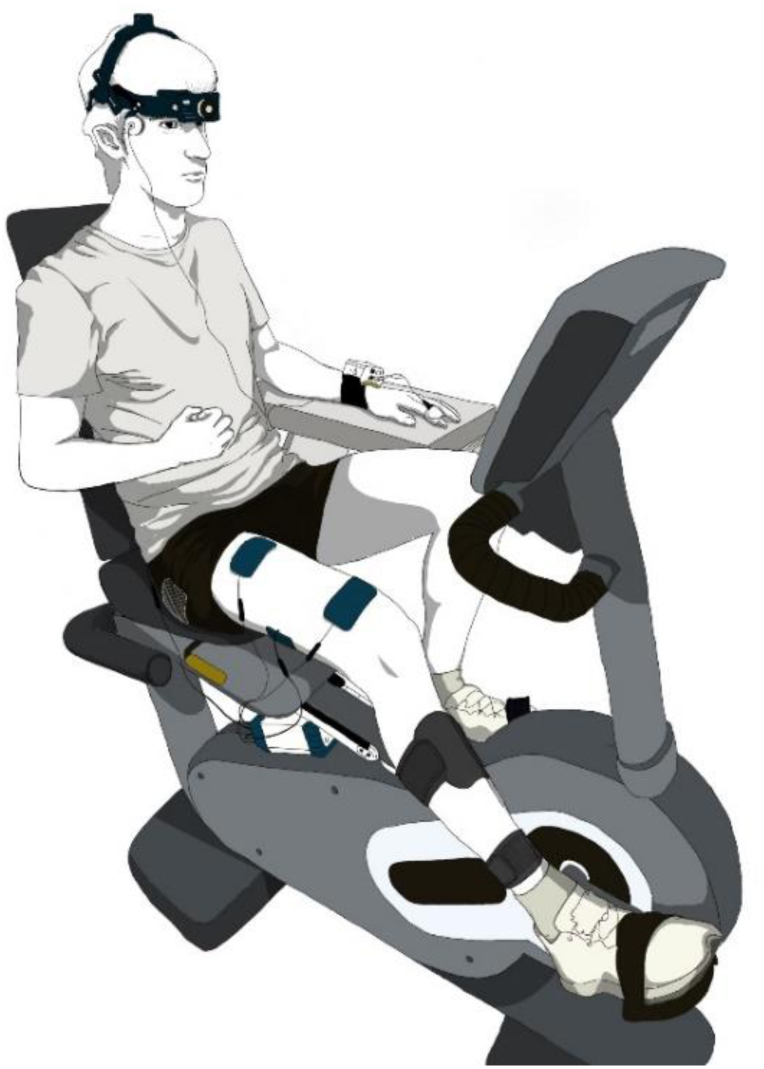
Experimental set-up. Electrical stimulation was applied to the paretic leg, on the quadriceps, hamstrings, and gluteus muscles.

### Measurements and data analysis

#### Perception of effort

The perception of effort is defined as “the conscious sensation of how hard, heavy, and strenuous the physical task is” (3,12). Participants reported the intensity of their perceived effort to cycle via the following question: ‘‘How hard is it for you to drive your legs?”. The intensity of the perceived effort was reported every 5 minutes during the cycling exercise, using the validated CR100 scale (13). The scale was presented to the patient on a laminated sheet, allowing them to report the intensity of their perceived effort in real time. This scale ranges from 0 for “nothing at all” to 100 for “maximal”. The scale contains verbal anchors such as light (13), moderate (23), strong (50), or very strong (70). The maximal intensity (100) was anchored based on previous experience and defined as “the maximal effort experienced in your life, for most people it corresponds to the most strenuous exercise previously experienced.” Patients were familiarized with the scale during the incremental test and the familiarization sessions (visits 2-3-4).

#### Cardio- and cerebrovascular parameters

Heart rate (HR) was recorded continuously using a heart rate monitor (Forerunner 630, Garmin, USA), mean arterial blood pressure (MAP = Diastolic Pressure + 1/3 (Systolic Pressure – Diastolic Pressure) and cardiac output (CO) were monitored using finger photoplethysmography attached to the non-dominant hand (Finometer PRO; Finapres Medical Systems, Amsterdam, The Netherlands). The blood flow velocity in the right middle cerebral artery (MCAv) was continuously monitored using a 2 MHz pulsed Doppler ultrasound system (TCD, Multi-Dop X4, DWL Elektroniche System, Singen, Germany). Nine patients volunteered for cerebral blood flow velocity monitoring. Each session began with confirmation of artery insonation, relying on expected signal depth, velocity, visual signal along, and carotid compression for validation. The Doppler probe was placed on the right transtemporal window and secured with a DiaMon headband (DWL) throughout all experimental sessions. The placement of the probe and the depth of the artery were carefully recorded to be the same for each condition.

Beat-to-beat data were continuously sampled at 1 kHz using an analog-to-digital converter (PowerLab/16SP ML795; AD Instruments, Oxford, UK) and stored for offline analysis on a computer (Chart version 7.2.2, AD Instruments, Oxford, UK). Cardio and cerebrovascular data were averaged for the last 30 seconds of each 5-minute interval period (before, during, and after the exercise) as done in a previous study (11).

#### Cognitive tasks

Cognitive tasks were conducted on a table and in a seated position, both before (PRE) and immediately after (POST) each experimental condition. The selected cognitive tasks included a Stroop Task and the Trail Making Tests (TMT-A and TMT-B), as performed in previous studies (11,14). Firstly, participants completed a three-phase Stroop task. These three phases included three versions of the Stroop tasks, referred to as the congruent, neutral, and incongruent conditions, respectively (11,14). Each task contained 100 items (words or crosses). On the first page, the words “blue,” “red,” “yellow,” and “green” were printed in black ink (congruent task – reading task). On the second page, “XXXX” was printed in blue, red, yellow, and green colors (neutral task). The last page displayed a color name written in a different font color from the printed word, and the participant had to name the ink color (incongruent task). Participants were instructed to read the words from left to right, aloud, as quickly and accurately as possible for 45 seconds. The number of words and errors was counted by the experimenter and recorded. Finally, these Stroop tasks were displayed on a 42-inch screen set to maximum brightness, with a standard distance of 150 cm between participants and the screen.

Lastly, the Trail Making Test (TMT) was performed. This test is separated into two parts. The Trail Making Test-A (TMT-A) consists of rapidly connecting a sequence of randomly placed numbers in ascending order from 1 to 25. The Trail Making Test-B (TMT-B) requires participants to alternately connect numbers and letters in sequence (e.g., 1 A, 2 B, 3 C, … 13). TMT performance was defined as the time (seconds) taken to complete the task. The test was performed with the patient’s unaffected hand due to spasticity in the affected hand.

#### Blood lactate

Given that blood lactate has been proposed as a potential key factor mediating the cognitive benefits of physical exercise (15,16), blood lactate concentration was measured. Blood lactate was collected using an automated analyzer (Lactate Pro, Scout 4), with a 5-μl blood sample collected from the tip of the non-paretic index finger (due to spasticity in the affected hand), at the beginning and 2 minutes after the end of exercise to assess peak concentration within an optimal window (17).

#### Patients’ experience with FES-assisted cycling – Qualitative data

Following both testing sessions, the researcher informed each patient that if they want, they are free to report to their experience with FES-assisted cycling and traditional cycling. No predefined questions or interview guide were used. With the patients’ explicit consent, their comments were manually and anonymously transcribed on paper, verbatim.

### Statistical analysis

Mean (SD). Statistical significance was determined at α < 0.05. The Shapiro–Wilk test confirmed data normality. Two-way repeated-measures ANOVAs (condition × time) were used. Post-hoc tests were adjusted for multiple comparisons using the Bonferroni correction. Effect sizes are reported with the partial eta square (partial η2), with values of 0.01, 0.06, and above 0.14 denoting small, medium, and large differences, respectively.

## Results

The average peak power output was 72 ± 36 watts, and the cycling exercises were performed at 36 ± 18 watts.

Changes in stimulation intensity during FES-assisted cycling, for each muscle, are available in the supplemental materials (Figure S1).

The perceived effort to cycle increased over time (time: *P* < 0.001; η_p_^2^ = 0.497) and was lower for FES-assisted cycling compared to traditional cycling (condition: *P* < 0.001; η_p_^2^ = 0.800) (Figure 2).

**Figure 2:**
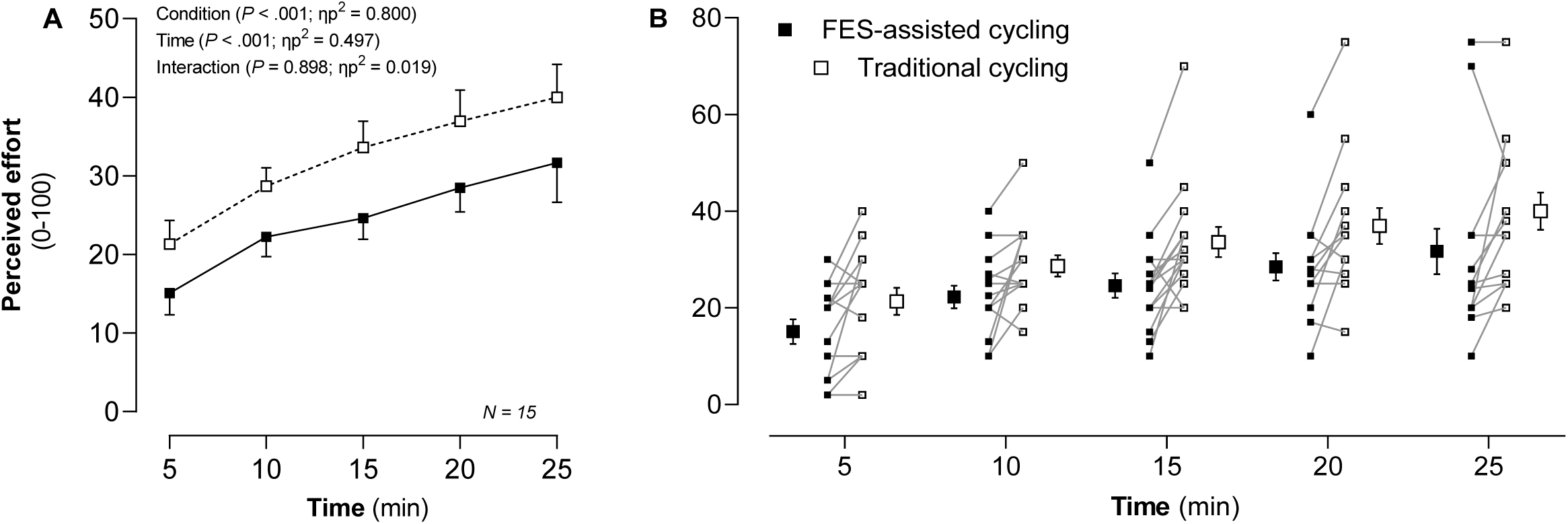
Perceived effort (A) and associated individual data (B) in response to FES-assisted (black) and traditional (white) cycling. Values are mean (SD). *n*, sample size for the analysis.

MCAv data is available for nine patients, as only these individuals volunteered for cerebral blood flow velocity monitoring. MCAv increases during both sessions (time: *P* = 0.026; η_p_^2^ = 0.407), with no difference between conditions (condition: *P* = 0.129; η_p_^2^ = 0.263) (Figure 3).

**Figure 3:**
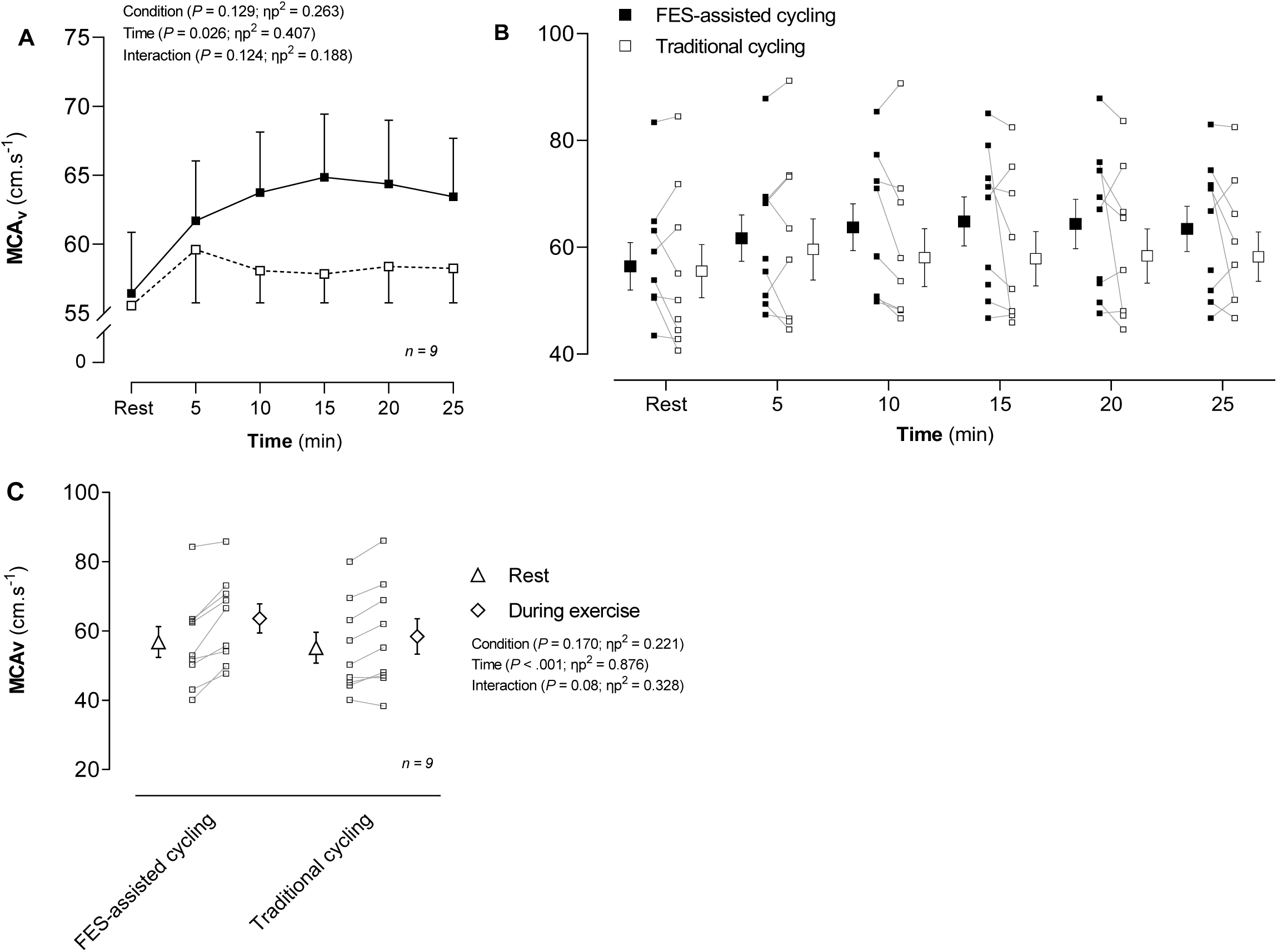
Middle cerebral artery velocity [MCAv (A)] and MCAv associated individual data (B), in response to FES-assisted (black) and traditional (white) cycling. (C) MCAv at rest (triangle), during exercise (rectangle), and associated individual data (white squares). Values are mean (SD). *n*, sample size for the analysis.

Due to technical issues relative to cable disconnection, PETCO₂ measurements were available for only 12 patients, and mean arterial pressure (MAP) and cardiac output (CO) for 7 patients. Blood lactate levels were available for 13 patients.

As shown in Figure 4, HR, mean arterial blood pressure (MAP), cardiac output (CO), PETCO_2,_ and blood lactate increased during exercise (Figure 3, all P < 0.001 and η_p_^2^ ≥ 0.327). Blood lactate levels were higher in the FES-assisted cycling session compared to the traditional cycling session (*P* = 0.018; η_p_^2^ = 0.384). CO and MAP are presented in supplemental materials—Figure S2 and increased in both sessions (all *P* ≤ 0.001 and η_p_^2^ ≥ 0.327).

**Figure 4:**
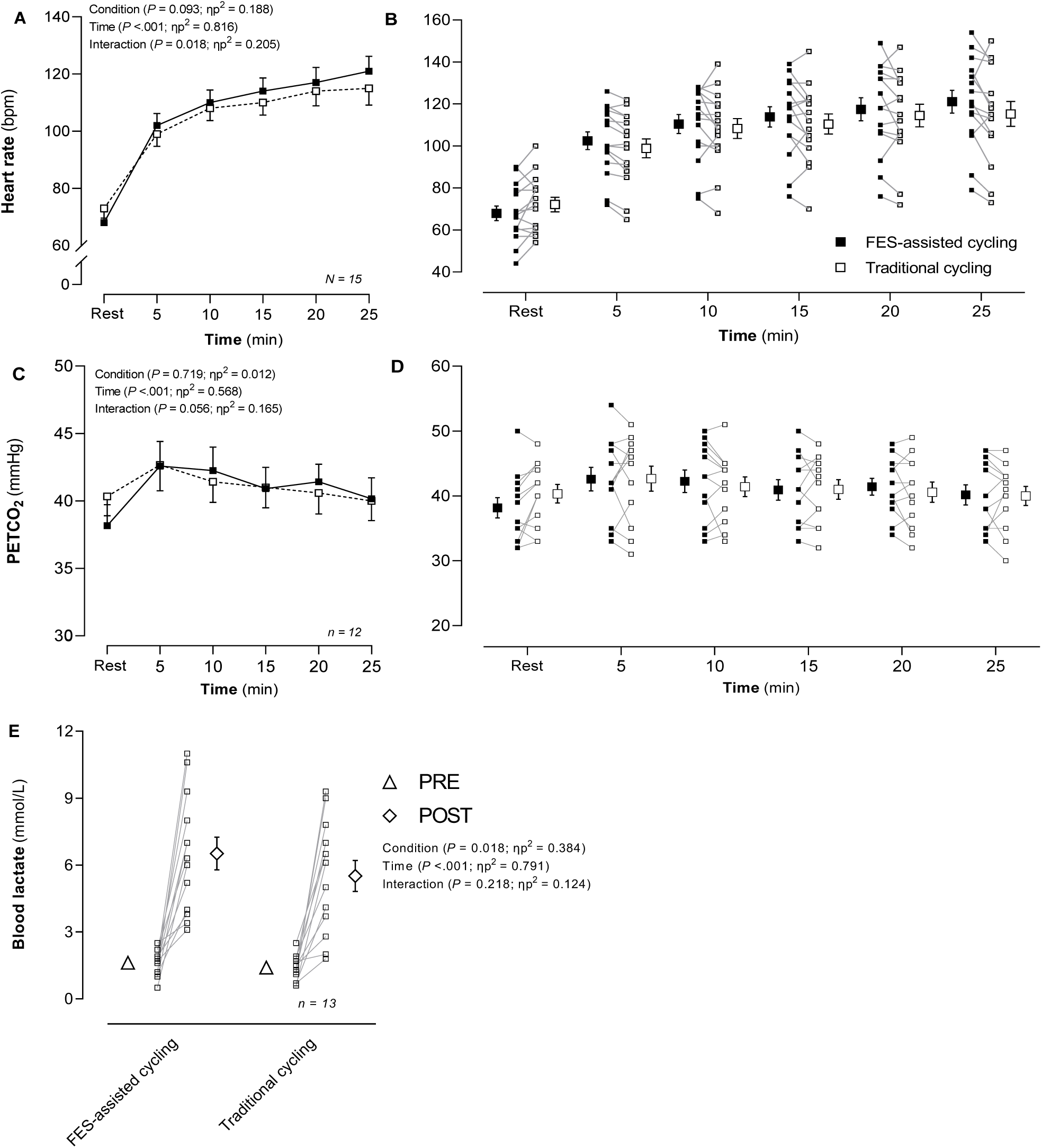
HR (A) and associated heart rate individual data (B), PETCO2 (C) and associated PETCO2 individual data (D), and blood lactate (E) in response to FES-assisted (black) and traditional (white) cycling. Data are presented as mean (SD). *n*, sample size for the analysis.

Due to fatigue or inability to complete the tasks, cognitive test data were available for 14 participants in the congruent Stroop condition, 13 in the neutral and incongruent conditions, and 12 for the TMT-B test. Details of the cognitive performance data is available in supplemental materials. Briefly, the results suggest that FES-assisted cycling induced similar effects of traditional cycling, with a potential positive effect specific to the Neutral Stroop task.

Eleven patients freely reported their experience with FES-assisted cycling. The verbatim of the experience reported is available in the supplemental materials—Table S1. The most recurring theme among the responses was a perceived benefit of the FES-assisted session compared to traditional cycling. Most of the participants reported that the FES made pedaling easier by inducing additional force during cycling, which reduced fatigue, and by improving coordination and fluidity of movement on the affected limb.

## Discussion

The increased perceived effort over the 20 minutes of FES-assisted and traditional cycling is caused by the progressive development of muscle fatigue, requiring an increased motor command to recruit additional motor units, leading to higher perceived effort levels (3,6,7). According to our hypothesis, when cycling at the same workload, the perceived effort was lower during FES-assisted compared to traditional cycling. To the best of our knowledge, this study is the first to observe in stroke patients the positive effect of FES-assisted cycling on the perception of effort. This result replicates previous observations in healthy individuals (18,19) and conceptually replicates previous observations of a lower perceived effort during a FES- walking exercise in multiple sclerosis (8) or stroke patients (9). It is well accepted that applying electrical stimulation to a muscle increases afferent feedback (20). Our results, combined with the studies observing a lower perception of effort during FES of the lower limb (i.e., cycling or walking), provide further evidence that the perception of effort scales with the magnitude of the motor command rather than afferent feedback (for review see (7)). However, the mechanisms underlying the FES-induced reduction in perceived effort remain to be fully understood and warrant further investigation. Brain imaging techniques would be of interest to better elucidate the relationship between the motor command and perceived effort during physical exercise involving electrical stimulation.

In our study, the lower perceived effort is likely linked to the force evoked by FES, which helped the patients by requiring only a small additional voluntary engagement to cycle. This interpretation is further supported by the patients’ experience qualitatively reported. Indeed, most of the patients reported perceiving clear assistance of the stimulation, helping the movement and facilitating the exercise. A limitation to this observation is that our study was not a-priori designed for a qualitative analysis. As our qualitative analysis to describe the patients experience with FES-assisted cycling was exploratory, we encourage future research to replicate our results by adopting a-priori a mixed-methods design. Doing so will allow a better interpretation on the relationship between quantitative measurement of effort and its perception, and the patients’ perspectives and experience. Moreover, we were unable to directly assess the actual contribution of FES to muscle force production and to measure the force output produced by the stimulator relative to the ergometer resistance. Thus, we cannot precisely quantify the extent to which FES contributes to force generation at different intensities. In the future, using force sensors attached directly to the pedals would allow for a more accurate assessment of this parameter, providing a better understanding of the role of FES in the perception of effort, in force generation, and its effective contribution to the cycling motion.

As a high perceived effort is a barrier to engagement in physical exercise and a key determinant of adherence to rehabilitation programs (3,21), the observed low perceived effort is of crucial interest for supporting the development and implementation of FES-assisted cycling in stroke patients’ rehabilitation. Future studies should now implement FES-assisted cycling during a rehabilitation program, and not only a single training session, to test the possibility that the effects of FES-assisted cycling on stroke patients’ rehabilitation could be superior to those induced by traditional cycling.

As cerebral blood flow has been proposed as one of the mechanisms underlying neuroplasticity (22), we also explored the CBF responses to FES-assisted cycling and its potential priming effect on cognitive performance in a subsample of voluntary patients. As previously observed during an isometric NMES session (11), our results suggest that FES-assisted cycling can increase CBF in the same proportions as cycling with lower perceived effort. These increases in blood velocity in the middle cerebral artery are not surprising, since similar results have been observed on pedaling tasks (23–26) and could be explained by the increases in cardiovascular parameters observed during exercise. As the literature has reported that the recruitment of a large muscle volume through electrical stimulation leads to a higher cardiorespiratory demand (5,27,28), we hypothesized that CBF would be higher in the FES condition. However, it was not the case in our study, and this could potentially be explained by the muscle volume stimulated in our FES condition. It is well established that the muscle mass engaged during physical exercise is a key factor influencing cerebral blood flow (29). Although a substantial muscle volume of paretic leg was stimulated in this study (quadriceps, hamstrings, and gluteal muscles), future research could explore the effects of stimulating an even greater number of muscles, including the entire lower limb (e.g., quadriceps, hamstrings, gluteus, tibialis anterior, etc.), rather than focusing solely on the paretic limb. Such an approach could enhance the effects of stimulation on CBF. However, further research is needed to optimize stimulation parameters and muscle selection, ensuring that potential adverse effects are avoided. One limitation of this study is that CBF was measured only on the right MCA, without distinguishing between the affected (ipsilateral) and unaffected (contralateral) sides. Thus, future research should consider measuring bilateral cerebral blood flow at the middle cerebral artery level to better understand the differences between the affected and unaffected sides.

Our results also suggest that cycling, whether assisted by FES or not, has limited and inconsistent effects on cognitive performance in stroke patients. Although both types of cycling sessions led to some improvements in cognitive performance, these effects were not consistently observed across all cognitive tasks (e.g., the incongruent condition of the Stroop task). These partial effects—improvements seen only in specific tasks—are consistent with the existing literature reporting mixed findings (10). For instance, recent studies have shown that an acute NMES protocol (11,14) can enhance cognitive performance, but only under certain conditions of the Stroop task in healthy individuals. Conversely, other studies have found no improvement in cognitive performance, particularly in tasks such as the Go/No-Go, among healthy participants (30,31). To address this discrepancy in the literature, we encourage future research to focus on the effect of FES-assisted cycling on cognitive performance as primary outcome of interest and design the study a-priori for this purpose.

Moreover, as shown in the present study, where elevated blood lactate levels were observed following FES, recent literature has reported that NMES applied to the quadriceps increases blood lactate levels in both humans and rats (14). Notably, higher blood lactate levels were associated with improved Stroop Task performance in healthy humans and increased hippocampal BDNF expression in rats following a single NMES session (14). These findings support the hypothesis that lactate may act as a key exerkine mediating the cognitive benefits of electrical stimulation. However, the underlying mechanisms remain poorly understood and warrant further investigation.

Lastly, we would like to remind the reader that our sample size was determined by our primary aim, i.e., testing the difference in perceived effort between FES-assisted cycling and traditional cycling. Due to the likely smallest effect size of FES-assisted cycling on CBF and cognitive performance, future studies should replicate our results with a specific a priori approach to CBF and cognitive performance and a higher sample size. Such studies could also unravel a potentially greater positive effect of FES-assisted cycling compared to traditional cycling, on CBF and cognitive performance.

To conclude, this study is the first to demonstrate a reduced perceived effort during FES-assisted cycling, compared to traditional cycling, in stroke patients. This study also suggests a potential positive effect of FES-assisted cycling on CBF, as observed in traditional cycling. Given that perceived effort is a significant barrier to engagement in and adherence to rehabilitation programs, this study highlights the strong potential of implementing FES-assisted cycling in stroke patients’ rehabilitation programs.

## Data Availability

All data produced in the present study are available upon reasonable request to the authors

## Declaration

### Ethics approval and consent to participate

The study was supported by UGECAM Rhône-Alpes and received ethical approval from the Comité de Protection des Personnes Sud-Est II (French Institutional Review Board). The study was approved under the clinical trial registration number NCT06230796 and registered at ClinicalTrials.gov. In addition, written informed consent was obtained from all subjects. Patients were assured that their information would only be used for analysis in this study. All methods were carried out under relevant guidelines and regulations.

### Consent for publication

Not applicable as all participants have been de-identified.

### Availability of data and materials

The data are available from the corresponding author on reasonable request.

### Authors’ contributions

**Conceptualization**, M.D, J.DM, J.V.B. & G.D; **methodology**, M.D, J.DM, J.V.B, B.P & G.D; **formal analysis**, M.D; **investigation**, M.D, J.DM & PH.P.; **data curation**, M.D; **writing—original draft preparation**, M.D, B.P & G.D; **writing—review and editing**, M.D, J.DM, E.J, J.V.B, B.P & G.D; **supervision**, M.D, J.DM, J.V.B & G.D; **figure designing**, M.D. & B.P. All authors have read and agreed to the published version of the manuscript.

## Acknowledgements

We thank Jean-Serein Mbende for co-designing the illustration included in this article.

## Supplementary Material/Additional file

### SUPPLEMENTRY FIGURE LEGENDS

Fig. S1. [Stimulation intensities (mA) used over the FES-assisted cycling modality.]

Fig. S2. [MAP (A) and associated individual data (B), CO (C) and associated individual data (D) in response to FES-assisted (black) and traditional (white) cycling. Data are presented as mean ± SE. *n*, sample size for the analysis.]

Fig. S3. [Cognitive performances in response to FES-assisted and traditional cycling. A) Congruent Stroop task; B) Neutral Stroop Task; C) Incongruent Stroop Task; D) TMT-A and E) TMT-B performances. PRE (triangle) and POST (square). Values are mean ± SE. n, sample size for the analysis. *: significant difference pre-post within the same condition.]

### SUPPLEMENTRY TABLE LEGENDS

Table SI. [Voluntary patient feedback following completion of both cycling sessions.]

