## Supplementary figures and images for "Positive effects of functional electrical stimulation-assisted cycling on perception of effort, cerebral blood flow and cognition in post-stroke patients"

### Additional figure 1

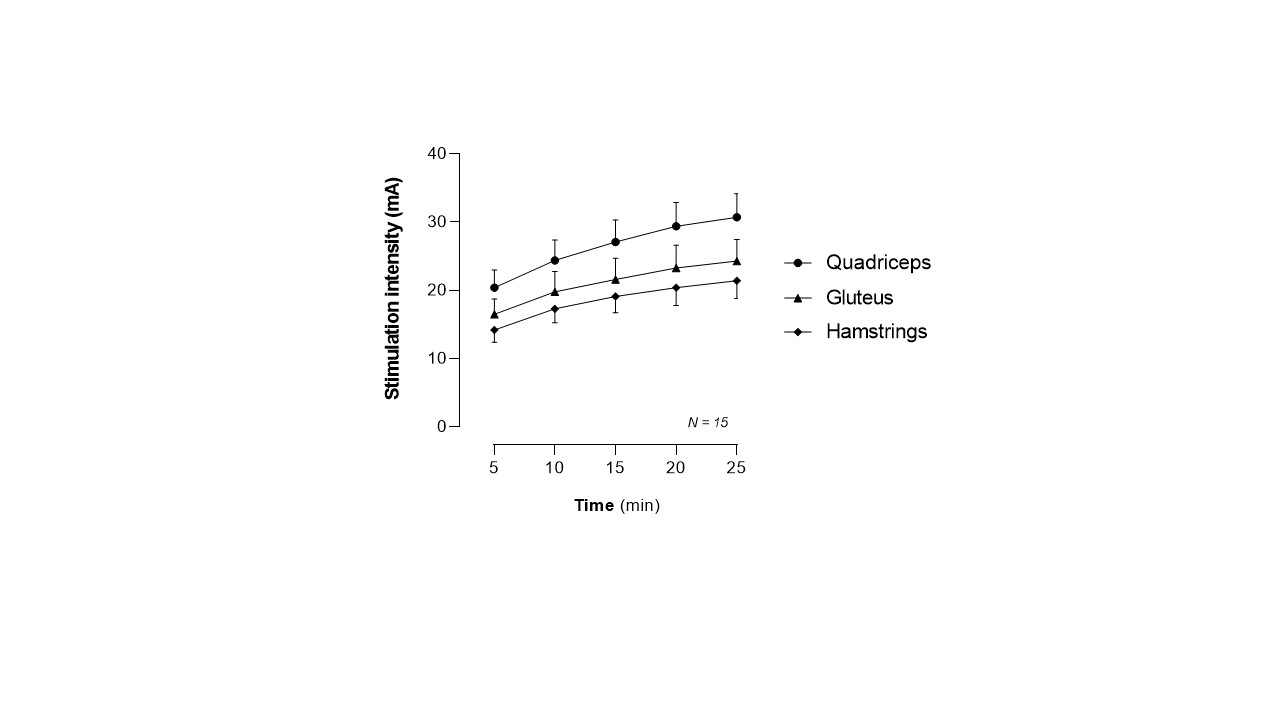

### Additional figure 2

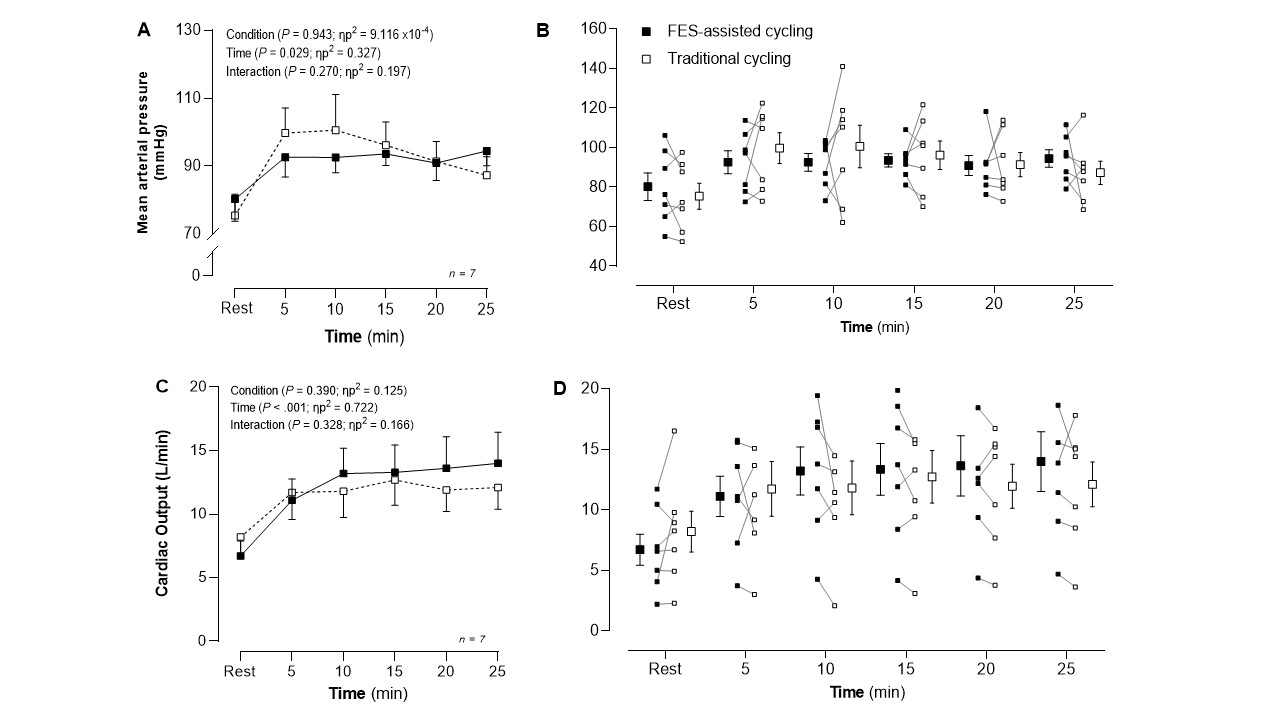

### Additional figure 3a

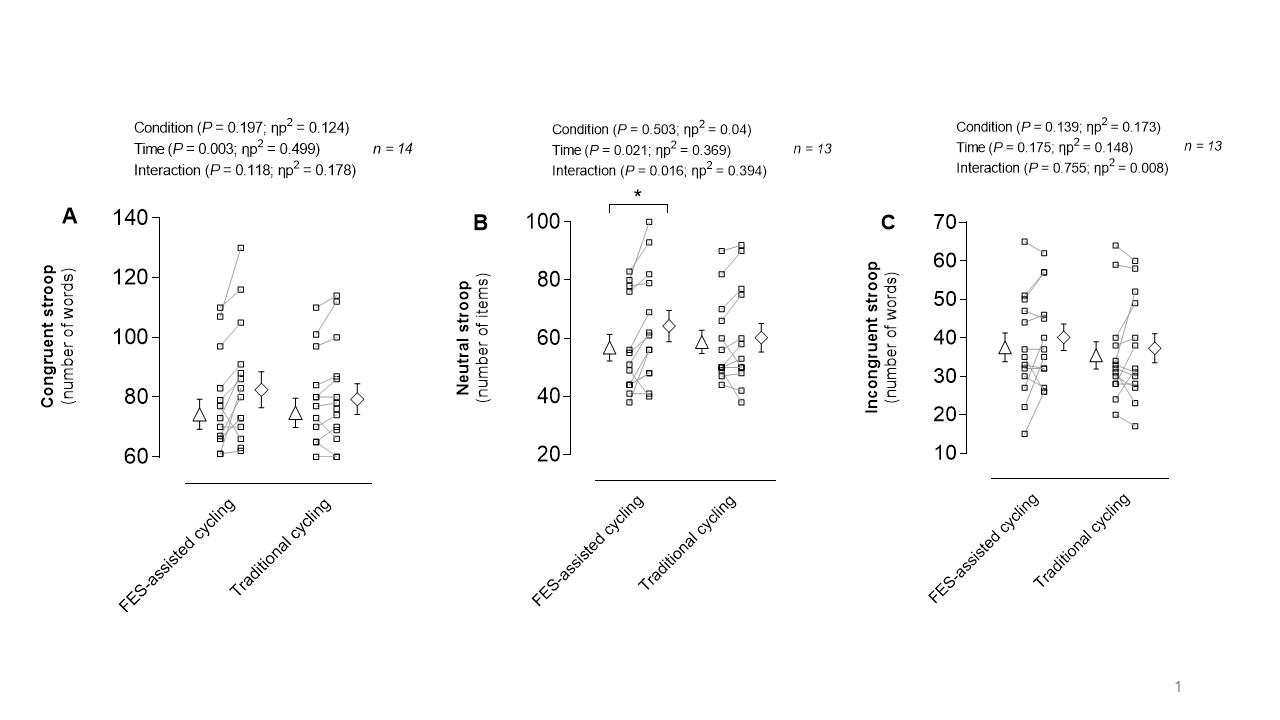

### Additional figure 3b

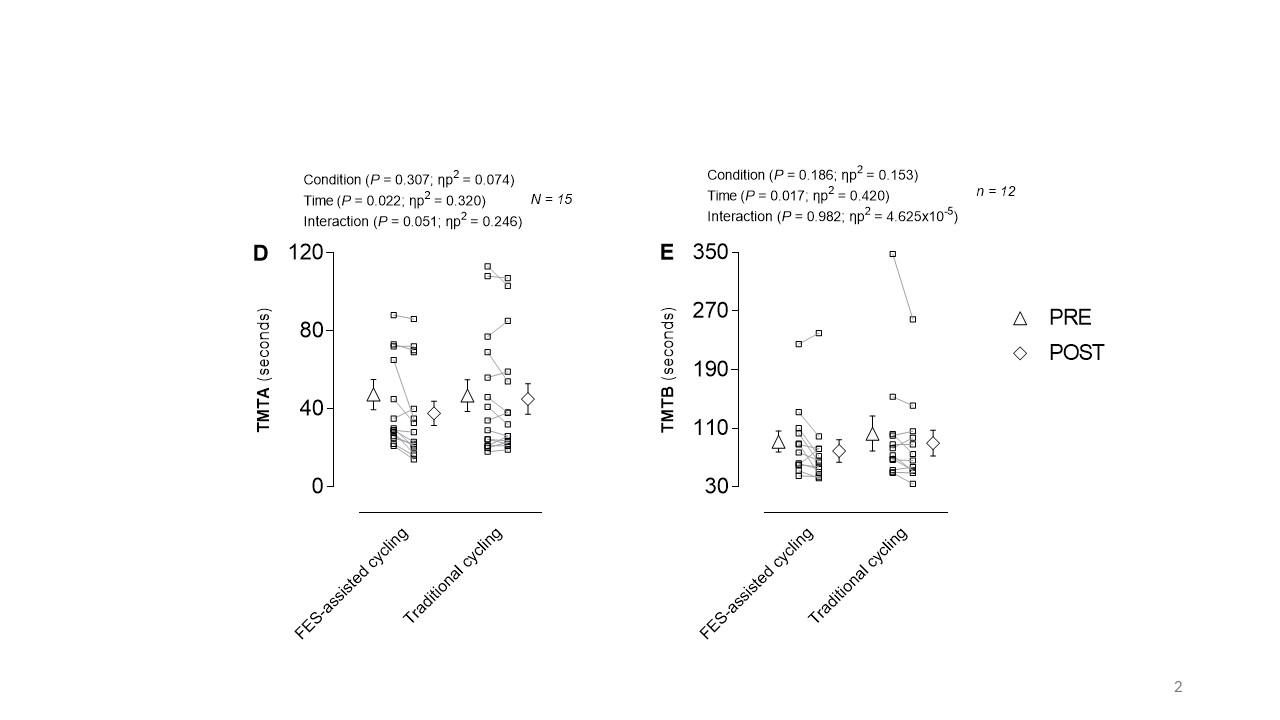
